# FDA and EMA clinical research guidelines: Assessment of trial design recommendations for pivotal psychiatric drug trials (Protocol)

**DOI:** 10.1101/2020.01.22.20018499

**Authors:** Kim Boesen, Peter C Gøtzsche, John PA Ioannidis

## Abstract

This is a protocol for the project entitled "FDA and EMA clinical research guidelines: Assessment of trial design recommendations for pivotal psychiatric drug trials".

## Background

### Uncertain patient-relevant benefits of new drugs

Several recent reports have found that newly authorised medicines often have questionable or no added patient-relevant benefits compared to already available, older treatments.^1-6^ In a cohort of 216 drugs approved by the German drug regulatory authority between 2011 and 2016, the independent Institute for Quality and Efficiency in Health Care (IQWiG) judged that 22 (10 %) new drugs had substantial benefits compared to already available treatments, while 125 (58 %) drugs had no proof of added benefits.^1^ Cohort studies of oncology drugs approved by the European Medicines Agency (EMA)^4^ and the US Food and Drug Administration (FDA)^5^ found that most drugs were approved without evidence of benefits on overall mortality or quality of life. Similarly, an analysis of oncology drugs approved by the EMA between 2014 and 2016 reported that many pivotal trials underpinning drug approvals were of high risk of bias due to problems with the trial design and or trial conduct.^6^ It has been argued that the threshold for new drug approvals has been lowered to accommodate the pharmaceutical industry’s interests,^7^ and researchers have advocated for more pragmatic, patient-relevant trials.^8^

### Field specific issues in psychiatry

Systematic reviews, particularly of antidepressants for depression^9, 10^ and of central stimulants for attention deficit hyperactivity disorder,^11-13^ have highlighted problems of low generalizability of psychiatric drug trials, similar to those identified in pivotal oncology drug trials. Common methodological limitations in these trials are small sample size, short trial duration, restricted trial populations in terms of allowed psychiatric comorbidity, risk of withdrawal effects due to previous exposure to the drug of interest, and the use of surrogates and rating scales rather than patient-relevant outcomes.^9-13^ In the German cohort of authorised drugs,^1^ one out of 18 newly approved drugs (combined psychiatry/neurology) was judged to have substantial added benefits compared to already available treatments.

### Drug regulatory agency guidelines on how to design pivotal trials

The FDA and EMA publish guidelines on how to design and conduct pivotal trials for new drug approvals. Whether there are important differences in the way FDA and EMA apply and enforce these regulatory guidelines is uncertain.

EMA describes their Clinical Efficacy and Safety Guidelines as follows: “*The European Medicines Agency’s scientific guidelines on the clinical efficacy and safety of human medicines help applicants prepare marketing authorisation applications. Guidelines reflect a harmonised approach of the EU Member States and the Agency on how to interpret and apply the requirements for the demonstration of quality, safety and efficacy set out in the Community directives*”^14^ and “*The Agency strongly encourages applicants and marketing*

*authorisation holders to follow these guidelines. Applicants need to justify deviations from guidelines fully in their applications at the time of submission. Before that, they should seek scientific advice, to discuss any proposed deviations during medicine development*.”^14^ The EMA research guidelines should therefore be understood as the minimal required standard for pivotal trial design within the European Union.

The FDA has also recently begun issuing guidelines for designing pivotal trials. The FDA describes these documents as, “*Guidance documents represent FDA’s current thinking on a topic. They do not create or confer any rights for or on any person and do not operate to bind FDA or the public. You can use an alternative approach if the approach satisfies the requirements of the applicable statutes and regulations*”.^15^

We want to systematically assess how these guidelines are developed and discuss the recommended trial designs from a clinical point of view.

## Research question

Question 1: Who are the committee members behind the research guidelines and what are their declared conflicts of interest?

Question 2. Who comment on the draft guidelines (stakeholders) and how much does each stakeholder contribute?

Question 3: How is the commenting phase on the draft guidelines organised?

Question 4. What trial designs are FDA and EMA recommending regarding four trial characteristics (see below) in pivotal psychiatric drug trials?

## Methods

### Drug regulator guidelines

#### Inclusion criteria

- Research guideline (draft or final version) published by the EMA or FDA on how to design pivotal trials for drug approval.
- Must contain guidance on all aspects of pivotal trial design. The guideline should not be specifically about one aspect of the trial design, such as inclusion criteria, outcome measures, or other specific aspects of trial design.
- The guidelines must cover psychiatric diagnoses as defined in current or previous versions of the *Diagnostics and Statistics Manual of Mental Disorders* (DSM) or the *International Classification of Diseases* (ICD).
- No restrictions on year of publication.
- We will include the newest published guideline, if more than one is identified on the same subject.

#### EMA Clinical Efficacy and Safety Guidelines

The EMA *Clinical Efficacy and Safety Guidelines* are available on EMA’s website. We will screen all guidelines archived under the tag “Nervous system” (https://www.ema.europa.eu/en/human-regulatory/research-development/scientific-guidelines/clinical-efficacy-safety/clinical-efficacy-safety-nervous-system).

*Preliminary results (1 October 2019): 12 guidelines; autism spectrum disorder, bipolar disorder, ADHD, OCD, depression, general anxiety disorder, panic disorder, social anxiety, schizophrenia, alcohol dependence, PTSD, and premenstrual dysphoric disorder*.

#### FDA Guidance Documents

FDA makes available their *Guidance Documents* on the FDA website (https://www.fda.gov/regulatory-information/search-fda-guidance-documents). We will screen all guidelines labelled with the tags: “drugs” (Product), “Clinical/Medical” (topic), and “Center for Drug Evaluation and Research” (FDA Organization).

*Preliminary results (1 October 2019): 132 entries of which 3 seemed relevant; antidepressants, ADHD, and alcoholism*.

### 1. Guideline committee members’ conflicts of interest

The FDA and EMA guideline committee members and their declared conflicts of interest are not available in the publicly available documents on the websites. We will therefore file Freedom of Information Act requests to access the committee member lists and their declared conflicts of interest. We have previously filed a request for access to the EMA ADHD guideline committee, which we were granted. We will report the proportion of committee members with declared conflicts of interest for each guideline.

### 2. Stakeholder comments’ documents

#### Inclusion criteria

- Must be an official document, or designated website, published by the EMA or the FDA dedicated to stakeholder comments on draft /concept papers of research guidelines.
- All comments in the document (or similar) will be included and counted.

#### EMA stakeholder comments

EMA makes available on their website dedicated documents with stakeholder comments on the regulatory draft guidelines. The comments and corresponding replies from EMA are collated in a single document and made available on the EMA website along the adopted research guidelines. We will assess all available stakeholder documents for the included research guidelines.

*Preliminary results (Oct 2019): 5 stakeholder comment documents available (ADHD, schizophrenia, alcohol dependence, PTSD and premenstrual dysphoric syndrome)*.

#### FDA stakeholder comments

*Preliminary results (Oct 2019): There are no comment documents linked to the guidance documents. There may be comments in meeting material from the Psychopharmacologic Drugs Advisory Committee (https://www.fda.gov/advisory-committees/human-drug-advisory-committees/psychopharmacologic-drugs-advisory-committee). We will identify the relevant minutes in case these are mentioned in the guidance documents*.

#### Stakeholder conflicts of interest

We will assess and categorise the stakeholders’ conflicts of interest. If the stakeholder is an association, academic society or similar, we will search for information related to the organisation’s funding and disclosed conflicts of interest for board or executive members. If the stakeholder is an individual, we will look up the person’s latest publications and extract the declared conflicts of interest. We will categorise the stakeholders as:

- **Industry**: Pharmaceutical companies and organisations.
- **Not-industry but with industry-related conflicts**: Organisation, associations or individuals with financial conflicts of interest to pharmaceutical companies.
- **Independent**: Organisations, associations and individuals without conflicts of interest to the pharmaceutical industry.
- **Unclear financial relationship**: There is insufficient information to categorise the stakeholder.

For each stakeholder comment document, we will quantify:

- Number of stakeholders commenting on each guideline
- Number of total comments on each guideline and number of comments from each stakeholder.
- Number of comments on each guideline separated into industry, non-industry and unclear financial relationship.

### 3. Commenting phase on the draft guidelines

We will explore how the commenting phases on the draft guidelines are organised. We will assess whether stakeholders are actively or passively recruited to comment on the draft guidelines, e.g. if specific stakeholders are actively invited through invitations. We will assess:

- How long the draft versions are available for commenting.
- Whether there are any restrictions on who is allowed to comment, e.g. whether specific credentials or affiliations are required.
- Is the commenting phase advertised, e.g. on social media, in newsletters, or in relevant journals to make people aware that the draft guidelines are open for input.

We will search the FDA and EMA websites for information and contact the regulators directly for additional information, if necessary.

### 4. Trial design recommendations

For all included research guidelines, we will extract the trial design recommendations regarding four specific trial characteristics:

- Duration of follow-up, also after the randomised phase ended
- Exclusion criteria related to psychiatric comorbidity
- Previous exposure to the drug, or drug class
- Efficacy outcomes (primary and secondary)

We will describe each design recommendation in the context of how the drug (or similar drugs) is being used, outcome preferences by the patients or established gold standard outcomes in the COMET database, and how the trial recommendations may impact the external validity. See table 1 for a full description of how we will assess each trial characteristic.

**Table 1.** Description of trial design recommendations.

| Trial design recommendation | Clinical context |
| --- | --- |
| Duration of follow-up | We will take into consideration how the drug, or similar drugs, is being used, i.e. for how long the drug is normally prescribed and used.<br>We will use observational or registry-based studies, such as public claims or insurance data, to compare clinical use with the recommended trial lengths. |
| Psychiatric comorbidity | We will consider whether psychiatric comorbidities are frequently associated with the investigated condition, and which psychiatric comorbidities are the most frequent.<br>We will base this on empirical data from patient samples of randomised trials, cross-sectional studies or surveys of the relevant patient-groups. |
| Previous exposure to the drug, or drug class | Including participants who have previously been exposed to the assessed drug and selecting them based on a positive response is called an “enriched design”. FDA describes the enriched design as a method to obtain an “enhanced benefit-risk relationship”,<sup>17</sup> by amplifying the beneficial effects and reducing the harms.<br>We will take into consideration the effects of stopping enrolled participants’ current treatment, often called a “washout”, before randomisation to drug or placebo. We will focus on whether a tapering or an abrupt stop is recommended, the drugs’ mechanisms of action, and the risks of developing abstinence symptoms.<br>We will search for empirical studies that have assessed the effects of enriched designs and “washouts” for the different drugs and conditions. |
| Efficacy outcome measures | We will search the Core Outcome Measures in Effectiveness Trials (COMET) database,<sup>19</sup> to see if a standardised set of outcome measures have been defined for the different conditions.<br>If not, we will use reports or surveys of patient preferences for the relevant conditions, such as Eiring et al.’s systematic review of patient preferences across mental disorders.<sup>20</sup> |

### Data extraction

One researcher (KB) will identify the research guidelines and stakeholder comments and extract outcome data. Another researcher will be involved when there is doubt about the inclusion of guidelines, stakeholder comments or data extraction.

### Data analysis

We will report:

- Total number of guideline committee members on each guideline, and proportion (%) of guideline committee members with declared conflicts of interest.
- Total number of stakeholders, total number of stakeholder comments on each guideline, and the proportion (%) of comments from each stakeholder category: industry, not-industry but with industry-related conflicts, independent, and unclear relationship.

We will not apply specific statistics on the analyses of the commenting phase and the description of the trial design recommendations.

## Discussion

### Strengths of the project

To our knowledge, this will be the first systematic and empirical assessment of the FDA and EMA guidelines on how to design pivotal trials. We have prespecified all outcomes in this protocol, which will be publicly available before the data extraction and data analysis begin. We believe it is an important question to address how two major drug regulators conceive their clinical research recommendations, if the guideline development is influenced by vested interests and whether the trial design recommendations may contribute to research waste.^16^

### Limitations of the project

The number of preliminary identified FDA Guidance Documents is small compared to the cohort of EMA guidelines. We also note that not all EMA guidelines have available stakeholder comments. We depend on the FDA and EMA on getting access to the lists of guideline committee members and their declared conflicts of interest, since these documents are not publicly available. If we are not granted access to these documents within a reasonable time frame, we may have to skip this research question.

### Timeline of the project

Data extraction and analysis will be conducted in January to March 2020. We expect to complete the project in early Spring 2020.

## Reporting and dissemination

This protocol will be made available on the medRxiv preprint server (https://www.medrxiv.org/). The final report and the corresponding dataset will be submitted for publication in a peer-reviewed medical journal.

## Data Availability

We will make available our full dataset on the Open Science Framework upon publication of the final report in a peer-reviewed medical journal. There may be restrictions on what we can share of documents obtained through the Freedom of Information Act requests.

## Acknowledgements

We thank the METRICS lab meeting participants for insightful comments on the draft protocol.

## Contributions

KB conceived the idea and wrote the first draft of the protocol. JPAI and PCG contributed to the design of the study. All authors critically revised the protocol. All authors approved the final version. KB is the guarantor.

## Data sharing statement

We will make available our full dataset on the *Open Science Framework* upon publication of the final report in a peer-reviewed medical journal. There may be restrictions on what we can share of documents obtained through the Freedom of Information Act requests.

